# Ammonium Sulfate Addition Reduces the Need for Guanidinium Isothiocyanate in the Denaturing Transport Medium Used for SARS-COV-2 RNA Detection

**DOI:** 10.1101/2022.02.28.22271591

**Authors:** Ge Liu, Jiaoyan Jia, Jianfeng Zhong, Hanfang Jiang, Yongqi Yang, Xiujing Lu, Zhendan He, Qinchang Zhu

**Author notes:** **Correspondence:** Xiujing Lu:; Zhendan He:; Qinchang Zhu.

## Abstract

Rapid identification of SARS-CoV-2 infected individuals through viral RNA detection followed by effective personal isolation remains the most effective way to prevent the spread of this virus. Large-scale RNA detection involves mass specimen collection and transportation. For biosafety reasons, denaturing viral transport medium has been extensively used during the pandemic. But the high concentrations of guanidinium isothiocyanate (GITC) in such media have raised issues around sufficient GITC supply and laboratory safety. Here, we tested whether supplementing media containing low concentrations of GITC with ammonium sulfate (AS) would affect the throat-swab detection of SARS-CoV-2 pseudovirus or a viral inactivation assay targeting both enveloped and non-enveloped viruses. Adding AS to the denaturing transport media reduced the need for high levels of GITC and improved SARS-COV-2 RNA detection without compromising virus inactivation.

## 1 Introduction

The ongoing COVID-19 pandemic has resulted in more than 422 million infections and 5.8 million deaths worldwide since its emergence in late 2019 [1, 2]. It appears to have now entered a new wave of infection sparking by the heavily mutated Omicron variant [3]. Although several vaccines (e.g., mRNA vaccines, inactivated vaccines, and viral vector vaccines) have been conditionally approved in some countries [4, 5] and various antiviral agents are in various stages of clinical trial [6, 7], there is currently no effective way to control the COVID-19 pandemic. Before all susceptible populations are fully protected, it remains extremely important to deal with the pandemic by controlling the infection sources and blocking the transmission routes, both of which rely on rapidly identifying SARS-CoV-2 infected individuals and isolating them from the population. Nucleic acid-based methods are the fastest methods for detecting SARS-CoV-2, especially in the early infection stages [8]. Such methods have played irreplaceable roles in the mass screening for SARS-CoV-infection. Large-scale nucleic acid testing has been implemented following local case reports where possible community transmission is indicated and this approach has supported China’s sustained containment of COVID-19 [9]. Wuhan (Hubei Province) performed city-wide mass screening using reverse-transcriptase quantitative polymerase chain reaction (RT-qPCR) testing for nearly 10 million people over a 10-day period [10], and Qingdao (Shandong Province) tested 10.9 million people for SARS-CoV-2 RNA in 5 days [11].

Large-scale nucleic acid testing involves large-scale specimen collection and transportation. Nasopharyngeal swabs and throat swabs are usually used in the specimen collection for SARS-CoV-2 RT-PCR testing [8]. After collection, specimens are generally preserved and transported in nondenaturing or denaturing media. Nondenaturing media, such as conventional viral transport medium and Amies transport medium, mainly contain saline buffer and antibiotics [12-14]. Nondenaturing media are used to maintain the integrity and infectivity of the viruses in the specimen. Therefore, nondenaturing media are used to detect viral nucleic acids and antigens or for culture-based viral detection. However, laboratory personnel risk infection when handling infectious specimens during the testing process, especially during large-scale testing [15]. Denaturing transport media usually contain guanidinium isothiocyanate (GITC) or other virus-inactivating denaturants, which is why they are only suitable for nucleic acid detection. One advantage of denaturing transport medium is its potential to reduce the risk of infection of laboratory personnel and improve SARS-CoV-2 RNA detection [16-18]. Both denaturing and nondenaturing viral transport media are widely used [13, 17, 19], and the former are recommended in China for large-scale nucleic acid testing [20]. Although various commercially available and in-house denaturing viral transport media exist, there are no uniform formulation standards. The core component of denaturing viral transport media is usually GITC, whose concentrations are mostly between 30%–50% (2.5M–4.2M) [17]. GITC is a chaotropic agent with a strong protein denaturing function. It is commonly used as a nucleic acid protector during RNA and DNA extraction because it denatures RNases and DNases. GITC is also used to deactivate many viruses including SARS-CoV-2 [17, 21, 22].

However, with the growing demand for denaturing viral transport media in large-scale nucleic acid testing, a GITC supply shortage occurred and its price doubled in early 2021 in China. Therefore, optimizing the formulation by reducing the GITC content may be a way to help meet the demand for SARS-CoV-2 nucleic acid screening and reduce the testing costs. Ammonium sulfate (AS) is a low-cost inorganic salt with high water solubility from its ionic nature. It is often used for protein precipitation and purification in the laboratory. High-concentration AS is also used to enhance RNA stability in tissue samples [23, 24]. Here, we report that reducing the amount of GITC in denaturing viral transport media while adding AS to them can improve the sensitivity of SARS-CoV-2 RNA detection without affecting the virus inactivation effect.

## 2 Materials and methods

### 2.1 Cells and viruses

Vero cells (ATCC, CCL-81) were cultured in Dulbecco’s modified Eagle’s medium (Gibco, NY, USA) supplemented with 10% (v/v) fetal calf serum. SARS-CoV-2 pseudovirus (FNV-2019-nCoV-abEN, >10^8^ copies/mL) was purchased from Fubio Biological Technology Co., Ltd. (Hangzhou, China). Herpes simplex virus type 1 (HSV-1, VR-733) originated from the American Type Culture Collection. Enterovirus 71 (EV71, C4 strain) was kindly provided by Dr Tao Peng, Guangzhou Medical University, China. Both HSV-1 and EV71 were propagated and titred in Vero cells. SARS-CoV-2 pseudovirus was used directly.

### 2.2 Denaturing solutions

Modified Primestore MTM (Molecular Transport Medium), which we used as a reference medium, contains 3M GITC, 25 mM sodium citrate, 0.5% SLS (sodium lauryl sulfate) and 20 mM ethylenediaminetetraacetic acid. Basing on the modified Primestore MTM, we prepared the denaturing transport medium with various GITC concentrations and/or AS as indicated.

### 2.3 SARS-CoV-2 RNA detection

A throat swab sample from a healthy person was collected in 4 mL saline, and the 100 μL of SARS-CoV-2 pseudovirus (>10^7^ copies) we added to the sample was mixed in by vortexing. Next, 200 μL of the SARS-CoV-2 pseudovirus-containing throat swab sample was added to 1 mL of each denaturing solution we prepared or to PBS. The solutions were kept at room temperature (RT) or 37°C, and RNA was extracted from them using the Virus DNA/RNA Kit (GBCBIO) after incubation for 24 h, 72 h, 120 h or 5 d. RTLJqPCR detection of SARS-CoV-2 RNA was performed in 20-μL reactions.The BeyoFast™ Probe One-Step qRT-PCR Kit (Beyotime) and primer and probe sequences targeting the ORF1ab gene and N gene were used in accordance with the protocols for COVID-19 Prevention and Control Guidelines (Seventh Version) issued by China’s National Health Commission. The cycle threshold (Ct value) for each sample was recorded and used for cross-sample comparisons. Graphs and statistical analyses (multiple t tests) were prepared with GraphPad Prism 8.0.2 (GraphPad Software, Inc, La Jolla, CA, United States). Statistical significance determined using the Holm-Sidak method, with alpha of 0.01.

### 2.4 Virus inactivation assay

The viral inactivation abilities of the denaturing solutions containing different GITC concentrations and/or AS were investigated using a plaque reduction assay. HSV-1 virus (100 µL, 4.135 × 10^8^ plaque forming units (PFUs)/mL) or EV71 virus (5 × 10^7^ PFUs/mL) was added to 0.9 mL of each denaturing solution, and each solution was mixed and incubated for 1 h at RT. After incubation, each mixture was serially diluted and filtered through Millex-GP 0.22 μm membrane filters (Millipore) and finally used in a plaque reduction assay using previously described procedures [25, 26].

## 3 Results and discussion

### 3.1 Effects of the test solutions on SARS-CoV-2 RNA detection

To evaluate the effect of adding AS to denaturing transport media on SARS-CoV-2 RNA detection, a modified commercial denaturing transport medium containing 3M GITC was used as the basic formula and reference reagent in the experiments. To mimic the sampling of SARS-CoV-2, a throat swab sample from a healthy person was mixed with SARS-CoV-2 pseudovirus. It was then placed into denaturing media containing different concentrations of GITC and AS and incubated at room temperature or 37°C for 24 h, 72 h, 120 h or 5 d as indicated. After RNA extraction, qRT-PCR on the SARS-CoV-2 ORF1ab and N genes was performed and comparatively analysed.

The results showed that reducing the GITC concentrations increased the Ct values, but adding 1M AS to the transport media reduced the Ct values of the ORF1ab gene and the N gene irrespective of the 24-h, 72-h or 120-h incubation period (Fig. 1A and 1B). A further assay on the samples incubated at RT or 37°C for 5 days revealed that adding 1M AS to transport media containing lower GITC concentrations significantly reduced the Ct values when compared with PBS or the transport medium containing 3M GITC (p<0.01) (Fig. 1C). A clear dose-dependent effect was observed when AS was added to each transport medium containing a fixed GITC concentration (Fig. 1D). Because lower Ct values indicate higher qRT-PCR detection efficiency, these results clearly suggest that adding AS to the transport media can reduce the amount of GITC required for sample denaturation and increase the detection efficiency for SARS-CoV-2 RNA.

**Figure 1.**
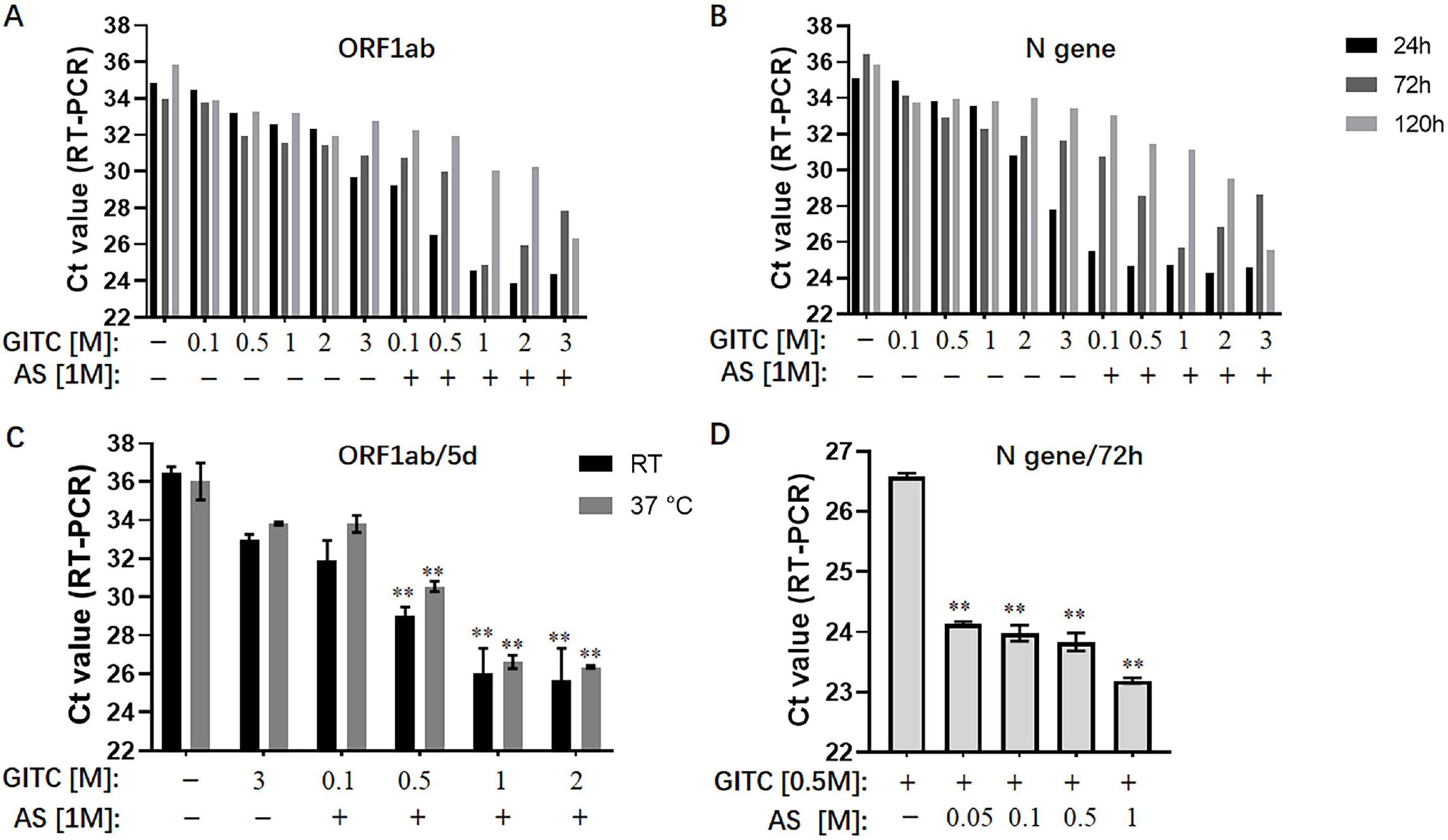
Effects of ammonium sulfate (AS) addition to denaturing transport media on SARS-CoV-2 RNA detection. (A) ORF1ab gene detection for various samples in denaturing media or PBS for 24 h, 72 h and 120 h at room temperature (RT). (B) N gene detection for various samples in denaturing media or PBS for 24 h, 72 h and 120 h at RT. (C) ORF1ab gene detection for various samples in denaturing media or PBS for 5 d at RT or 37°C.**p<0.01 vs. 3M GITC group. (D) N gene detection for various samples in denaturing media across an AS concentration gradient for 72 h at RT. **p<0.01 vs. Control group. GITC: guanidinium isothiocyanate. Samples without GITC and AS are the PBS controls.

GITC is commonly used at high concentrations in denaturing viral transport media [17, 21]. However, the sharp increase in demand for large-scale nucleic acid testing is causing a shortage of GITC during the ongoing COVID-19 pandemic. Another issue is that the high GITC concentrations used in the viral testing platforms can react with bleach (sodium hypochlorite) to produce harmful cyanide gas [27]. Therefore, reducing the GITC concentrations in transport media is worthwhile. However, we found that simply reducing its concentration in transport medium decreased the detection efficiency towards SARS-CoV-2 RNA (Fig. 1A and 1B). Here, we showed that adding AS to the transport media not only recovered but also improved the detection efficiency when low concentrations of GITC were present. The use of AS to precipitate proteins out of solution is known, and AS have been reported to suppress more than 90% of RNase A activity [28, 29]. Therefore, the improved RNA detection efficiency observed in this study possibly resulted from AS-induced denaturation of the RNase A protein, but further research is needed to confirm it.

### 3.2 Inactivation effects on enveloped and non-enveloped viruses

To compare the virus inactivation effects of denaturing transport media containing low concentrations of GITC and 1M AS with a commonly used denaturing transport medium, a plaque reduction assay designed for enveloped and non-enveloped viruses was performed. SARS-CoV-2 is an enveloped virus. However, the biosafety hazards relating to conducting experiments with a live SARS-CoV-2 virus caused us to use a common enveloped virus, HSV-1, to evaluate the inactivation effect of various transport media against this virus. We also tested EV71, a non-enveloped virus with a typical icosahedral capsid structure. After subjecting the viruses to various denaturing solutions at RT for 1 h, each virus was subjected to a plaque reduction assay.

The results showed that a common denaturing viral transport medium containing 3M GITC completely inactivated both HSV-1 and EV71 (Fig. 2A and 2B). No difference was observed in HSV-1 inactivation when 1M AS was added to the media even when the GITC concentrations were reduced to 0.1M (Fig. 2A). In the EV71 inactivation experiments, the effect of supplementing the transport media with 0.5 M GITC plus 1M AS was equal to that where 3M GITC was present, and EV71 was completely inactivated (Fig 2B). These results suggest that decreasing the GITC concentrations but adding AS to the transport media did not affect the virus inactivation effect when compared with general denaturing transport medium containing GITC at high concentration.

**Figure 2.**
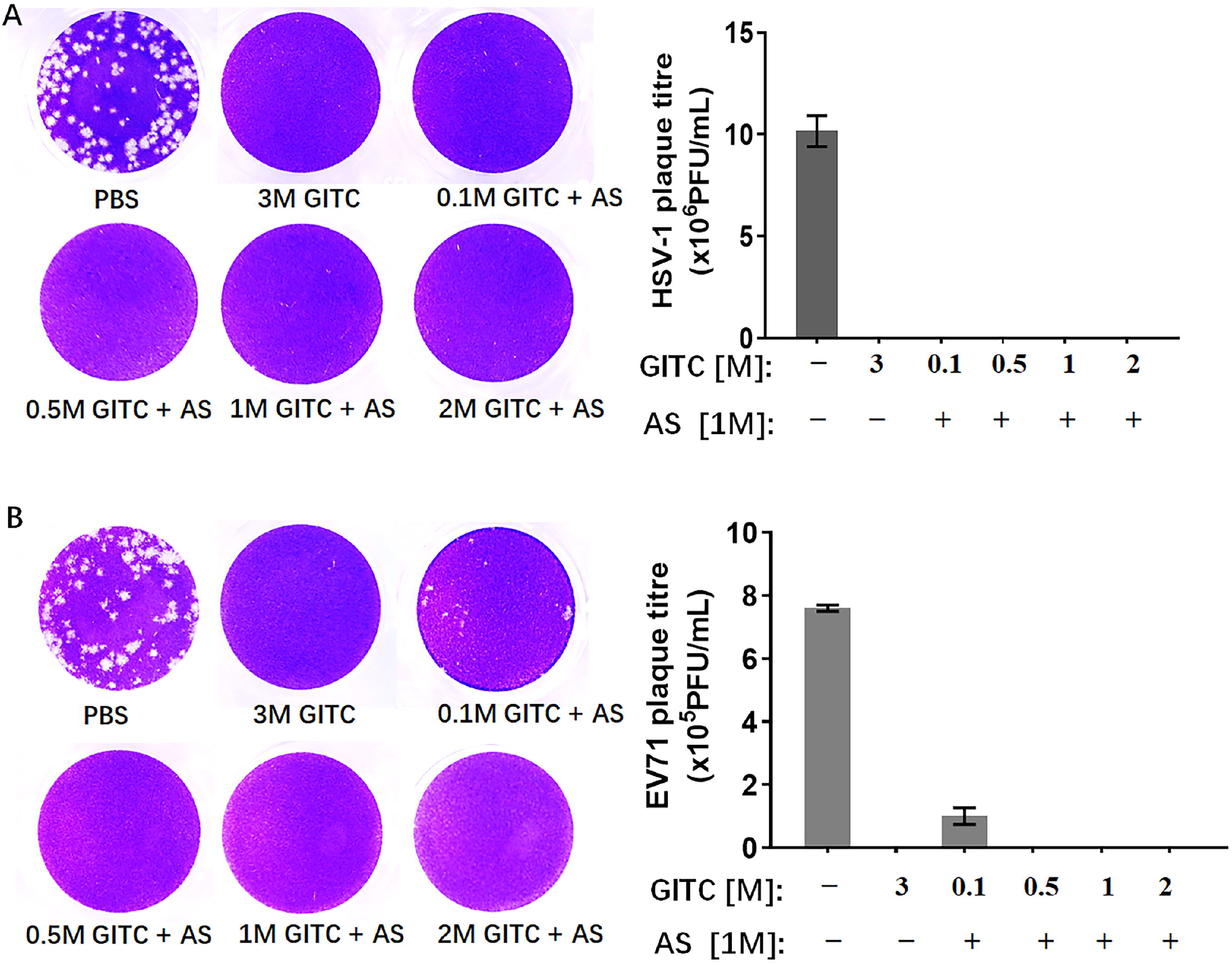
Effects of denaturing transport media containing low concentrations of guanidinium isothiocyanate (GITC) and ammonium sulfate supplementation on virus inactivation. (A) Plaque assay targeting the enveloped HSV-1 virus. (B) Plaque assay targeting the EV71 non-enveloped virus. Viruses were incubated in PBS or transport media containing the indicated components, followed by plaque assays on Vero cells. Plaques were visualized by crystal violet staining. Left panels: representative plaques. Right bar charts: virus titres calculated from the plaque assays. Data represent the average ± SD from three independent experiments.

## 4 Conclusions

Faced with the ongoing global COVID-19 pandemic, we sought a safer, low-cost denaturing viral transport medium for preserving SARS-CoV-2 RNA without compromising the effectiveness of viral detection. Based on the components of the modified commercial transport medium we used as a reference reagent, we reduced the GITC concentrations in the media, added AS, and obtained a new formula. We tested the different transport media by detecting SARS-CoV-2 pseudovirus in a throat swab sample and via a viral inactivation assay targeting both enveloped and non-enveloped viruses. We found that adding AS to the denaturing transport media reduced the use of GITC, improved SARS-COV-2 RNA detection, and did not compromise the virus inactivation effect of the media. These findings suggest that AS is a potential component that when added to denaturing transport medium may reduce the cost and improve the detection efficiency towards SARS-COV-2 RNA.

## Data Availability

All data produced in the present work are contained in the manuscript

## Author Contributions

LG conceived, planned the experiments and wrote the manuscript. JJY, ZJF and YYQ carried out the experiment. JHF analyzed the data and reviewed the manuscript. LXJ, HZD and ZQC conceived the original idea, acquired financial support and supervised the project.

## Acknowledgments

We thank Professor Tao Peng of Guangzhou Medical University for providing enterovirus 71. We also thank Dr. Sandra Cheesman for editing the language of a draft of this manuscript.

